# Community variability in TB-related stigma in South Africa: an ecologic analysis from the MISSED TB Outcomes Study

**DOI:** 10.1101/2025.04.15.25325605

**Authors:** Aaron M. Kipp, Dawie Olivier, Nelisiwe Skonje, Liyabuya Majiza, Emilyn Free, Kristopher J. Preacher, Amrita Daftary, Nondumiso Ngcelwane, Andrew Medina-Marino

## Abstract

**Introduction:** Tuberculosis (TB) stigma is a critical barrier to timely diagnosis and treatment. Although stigma originates within communities, few studies have quantified community-level TB stigma or its variability across geographic contexts. This study describes methods for capturing community-level TB stigma and examines stigma variability across 93 urban, peri-urban, and rural communities in Buffalo City Metropolitan Health District, South Africa.

**Methods:** As part of the MISSED TB Outcomes Study, heads of household (HoHs) were surveyed in a geographically clustered random sample of households across demarcated study communities. Validated scales were used to measure perceived community-level TB stigma, HIV stigma, and TB/HIV knowledge. Demographic data, including self-reported household TB and HIV history, were also captured. Community-level data, including TB and HIV stigma, were generated by aggregating individual responses within each study community. Associations between TB stigma and other community-level variables were analyzed using robust linear regression.

**Results:** Surveys were completed by 3,869 households across 93 communities. Median community TB stigma scores varied significantly by community location, with rural communities reporting the lowest stigma and peri-urban communities the highest. TB stigma was positively associated with HIV stigma across all community types, with the strongest associations in urban and rural communities. No associations were observed between TB stigma and TB prevalence, TB knowledge, or household demographics after adjusting for community location.

**Conclusions:** TB stigma varied meaningfully across communities and was influenced by urbanicity and HIV stigma. These findings suggest that stigma-reduction interventions must be tailored to local contexts and consider community-level determinants beyond individual knowledge or TB burden. The identified variability in TB stigma will inform future multilevel analyses of the TB care cascade in South Africa.

**KEY MESSAGES:** *What is already known on this topic:* Studies of TB stigma primarily focus on individual-level factors that determine stigma. There is very little known about community-level TB stigma and potential determinants of its variation across communities.

*What this study adds:* This study found substantial variability in TB stigma across 93 study communities. Urban/rural status and community-level HIV stigma were important factors, while community TB prevalence and TB knowledge did not appear to be associated with stigma.

*How this study might affect research, practice or policy:* Our on-going research aims to identify additional community-level drivers that can inform TB stigma interventions. Urban versus rural status is likely a proxy for other drivers of stigma.

## INTRODUCTION

The World Health Organization estimates that in 2023 10.8 million people globally developed TB disease and 1.25 million died.^1^ That same year, South Africa’s estimated TB incidence was 427 cases per 100,000 population^2^, one of the highest in the world, and remains one of the high-burden countries for HIV-associated and drug-resistant TB.^1^ Yet South Africa is also the only high-burden country to have reduced TB incidence by 50% compared to 2015 rates, meeting the 2025 target of End TB Strategy (ETBS).^1^

Historically, national TB programs have focused on treatment success rates, which do not account for upstream indicators (e.g., case finding; treatment initiation). This was powerfully evident from an analysis of the South African TB care cascade, which estimated that 23% of cases were missed, not diagnosed, or never initiated treatment.^3^ When including the 15% of cases not successfully completing treatment, nearly 40% of those with TB in South Africa were not successfully identified, cared for, and treated. While a TB care cascade framework can identify major attrition points for intervention^4, 5^, this approach seldom identifies specific risk factors contributing to patient attrition or delays between successive steps. Furthermore, while individual- and system-level factors impacting progression along the TB care cascade have been identified^6–9^, community-level risk factors potentially amenable to intervention (e.g., TB and HIV stigma) are rarely investigated.

Tuberculosis stigma remains a major concern for detecting and successfully treating people with TB.^10^ Stigma has been broadly defined as the convergence of “labelling, stereotyping, separation, status loss, and discrimination occurring in a power situation that allows them to unfold”.^11^ Specifically, this results in a separation of “Us” versus “Them” by those who have the power to do so. As a result of this social process, those who have a specific health condition may experience “exclusion, rejection, blame, or devaluation that results from experience or reasonable anticipation of an adverse social judgement” because of their condition, even when such judgement is not medically warranted.^12^ In this way, societal attitudes towards individuals with a particular health condition can be manifested at the structural, community, and/or institutional level. The effects of these attitudes are experienced at the individual level as anticipated stigma (concern about prejudice, discrimination, or devaluation from community members), experienced stigma (discrimination, rejection, or isolation due to TB status), and/or internalized stigma (endorsing negative attitudes such as shame or blame about oneself due to one’s TB status).^10^

Although there is widespread recognition that much of the stigma towards people with TB originates within communities, most stigma studies (qualitative and quantitative) reduce the focus to individual-level factors. Many qualitative studies from Sub-Saharan Africa report TB stigma, driven by poor TB knowledge, fear of infection, connection with HIV, and drug-resistant TB, as a barrier to progression along the TB care cascade.^13–23^ Quantitative studies of TB stigma and delay in testing uptake are less common in Sub-Saharan Africa and report contradictory results.^24–36^ These studies are primarily cross-sectional and clinic-based and therefore miss those who never present for testing. Quantitative studies of TB stigma and treatment adherence or completion are not common, particularly in Sub-Saharan Africa.^25, 37–46^ These studies are generally prospective and report TB stigma to be associated with lower adherence and worse treatment outcomes, but few utilized validated stigma measures. Even fewer evaluated different TB stigma domains, but these suggest stigma may differentially impact various steps in the care cascade. ^25, 26, 39^

There have been limited efforts to intervene on TB stigma. The few interventions implemented focus on individual-level changes, following the trend of observational studies discussed above, and neglect attending to social structures which help shape, impact and intersect with TB stigma, (e.g., HIV-stigma, gender-norms, poverty).^47, 48^ To design and implement effective stigma interventions, its multilevel and intersecting drivers and impacts must be understood.^10^ Such insights can then inform the development of community-level interventions to promote individual TB care seeking behavior. Thus far, few studies report variability in TB stigma across districts, regions, or countries in Sub-Saharan.^49–53^ While rural/urban differences in TB stigma have been identified, there have been almost no attempts to investigate community-level factors that could explain this variability. One exception is Rood et al. who report individual- and country-level factors associated with TB stigma (wanting to keep a family member’s TB a secret) using country Demographic Health Survey data.^50^ They found that a higher national TB incidence was associated with lower TB stigma, while a higher national prevalence of both TB transmission knowledge and HIV were associated with higher TB stigma.

A better understanding is needed of how and to what extent community-level TB stigma and other social determinants impact outcomes along the TB care continuum, especially in the context of high HIV burden. Without targeted interventions and resources to address these issues, South Africa will not remain on track to reach its post-2025 ETBS targets. To address these critical gaps in knowledge, we conducted a prospective, mixed-methods study on multilevel and intersectional stigma and other social determinants affecting TB detection, care, and treatment (the MISSED TB Outcomes study). The goal of this paper is to describe the community-level data collection methods of the MISSED TB Outcomes study, describe the variability in TB stigma levels across study communities, and explore community-level socio-demographic and TB-related correlates of higher community-level TB stigma.

## METHODS

### Study location and population

This study was conducted in Buffalo City Metropolitan (BCM) Health District, Eastern Cape Province, South Africa (See supplemental figure S1). Eastern Cape Province has the highest TB notification rate in South Africa (703 per 100,000 population) with BCM having one of the highest notification rates in Eastern Cape Province.^3^ Among people with TB initiating treatment, BCM has lower treatment success (74.1%) than that of the province (76.6%) and country (77.9%).^54^ Based on annual TB headcounts and in consultation with the BCM Department of Health, 18 primary health care clinics located in urban, peri-urban, and rural areas were selected for study inclusion. From February through July 2022, Google Earth satellite images were used to demarcate clinic catchment areas. These were then divided into smaller demarcated study communities using South African enumeration areas and natural boundaries (e.g., roads, gullies or natural land breaks) in consultation with community members’ understanding of communities and neighborhoods. In total, 93 study communities were demarcated (46 urban, 26 peri-urban, and 21 rural) (See Figure 1, upper panels, for example).

**Figure 1.**
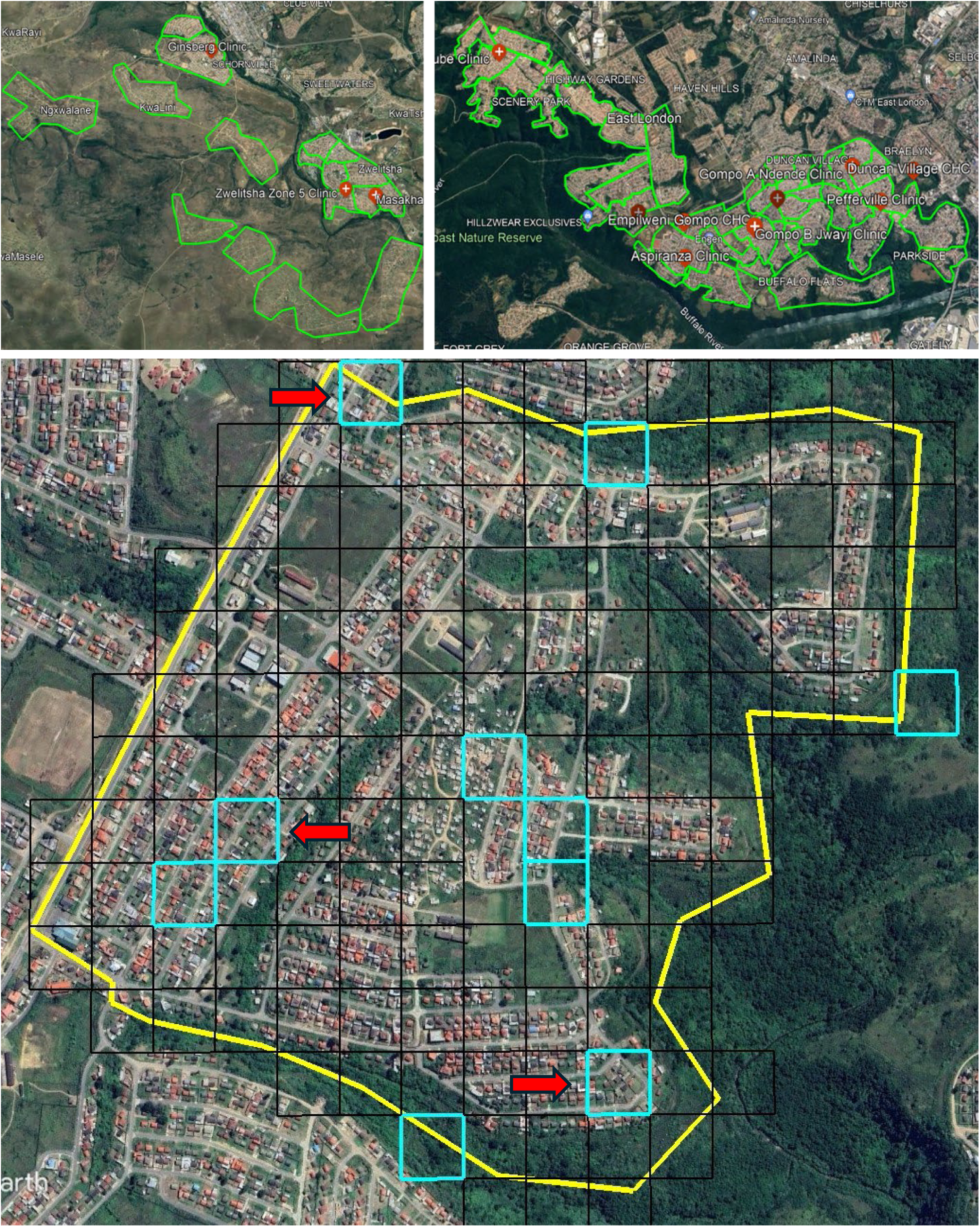
Example of demarcated study communities (green shapes) in rural (upper left panel) and urban (upper right panel) areas. Bottom panel illustrates geographic cluster sampling using a community <u>not</u> <u>involved in the study</u>. Blue boxes indicate the 10 randomly selected squares from the grid overly; Red arrows identify the first 3 squares to be approached, with remaining squares serving as supplemental if needed.

### Household sampling

For each of the 93 study communities, households within the community were selected for surveying using a single-stage geographic cluster sampling design.^55, 56^ To balance field logistics with the representativeness of the sample, we set the target sample size as a minimum of three separate areas within each community that could provide 8-12 participating households, for a total of 24-36 participating households per study community. To achieve this, community outlines from Google Earth were imported into a free, open-source mapping software (QGIS version 3.24.1; https://qgis.org/) that included the Google Satellite Hybrid plugin. QGIS was used to overlay each demarcated community satellite image with an initial 50m x 50m or 100m × 100m grid, depending on the approximate housing density in the community. Initial review of the grid was done to ensure the grid size generally allowed for 15-20 households per square (to allow for household non-response or vacant structures). As needed, a larger 150m x 150m grid size was used in rural areas, while smaller 25m x 25m grid were used in some dense urban communities. After selecting the final grid size for each study community, any squares that did not lie on or within the community boundary were discarded. The final gridded community files, which included a unique identification number for each square in the grid, were exported back to Google Earth for further labelling of sampled clusters.

For each community, 10 squares were randomly selected from the grid. See Figure 1 (bottom panel) for a hypothetical example from a community not involved in our study. The first three squares were the primary squares for household surveying. The remaining seven squares were used as supplemental squares if a minimum of 8 households per square was not met. Supplemental squares would be needed if a primary square fell close to the boundary of the community, on an open or commercial area, or a substantial number of the households were not able to be enrolled. When utilized, all households in a supplemental square were approached. If a selected square (primary or supplemental) appeared to have >25 households, it was sub-divided into four quadrants, randomly labelled A through D. If the sub-divided square was used, all households in the ‘A’ quadrant were first approached. Subsequent quadrants were used if the ‘A’ quadrant was not sufficient to achieve the minimum requirement of 8 households. The Google Earth satellite image, community outline, and outlines and labels of the 10 selected clusters were exported as a high-resolution image for use by the field survey teams.

The single-stage, geographic cluster sampling process was utilized as a pragmatic approach to balance the need for representative sampling with the logistical challenges of working in both urban and rural settings where reliable enumeration of households is not available; the most recent South African Census was conducted in 2010, and many of our study communities include dynamically changing informal housing and settlements.

### Data collection

All households whose front door fell on the boundary or within a square (or quadrant if sub-divided) were approached by trained field workers. Heads of households (HoH), age ≥18 years old (male or female), were identified by members of the household. Up to three attempts were made to meet with the HoH, including at least once on a weekend. HoHs were told about the study, and those interested in participating were consented by study staff. Each HoH completed a comprehensive, self-administered or interviewer-administered study questionnaire in either English or Xhosa using REDCap.^57, 58^ The survey included questions about the HoH and household demographics and previous history of TB, the HoH’s TB and HIV knowledge, and validated measures of TB and HIV stigma. Participants were compensated for their time with a small food voucher.

If the target enrolment for a particular square (≥8 surveyed households) was not achieved, the study team moved to the supplemental square and repeated the process, ensuring all possible households in that supplemental square were approached. Enrollment occurred from June 2022 through January 2023.

### Data analysis

The primary variable of interest was community-level TB stigma. HoHs completed a 10-item scale on perceived community stigma towards TB (e.g., “People may not want to eat or drink with friends who have TB”; “People are afraid of those with TB”) that had been previously validated in South Africa and has been used extensively in Sub-Saharan Africa.^52, 59–61^ Each item had a four-point Likert-type response option from strongly disagree (coded as 0) to strongly agree (coded as 3). Responses were summed and divided by the number of items to create an individual HoH TB stigma score ranging from 0 to 3, where higher scores indicated higher perceived community stigma. For each study community, HoH scores were averaged to create a composite community-level TB stigma score.

Other TB-related measures included TB knowledge and self-reported household TB prevalence. TB knowledge was measured using 18 items asking about the cause, transmissibility, symptoms, and treatment of TB.^62^ One point was assigned for each correct response, resulting in a TB knowledge score ranging from 0 to 18. As with TB stigma, these scores were averaged across HoH respondents to create a composite community-level TB knowledge score. Household TB prevalence was calculated as the proportion of surveyed households in the community who reported at least one household member previously being diagnosed with TB. Perceived HIV stigma was measured using 11 items developed and validated for use in South Africa.^63^ HIV knowledge was measured with an 18-item index previously used in Sub-Saharan Africa.^64–66^ Scoring of the HIV measures and creation of community-level scores were done in the same way as the TB stigma and knowledge measures.

Demographic characteristics of each HoH respondent were similarly averaged across all participating households in each study community to create the following community-level variables: 1) mean HoH age, 2) proportion of HoH who were female, 3) proportion of HoH who were Black South Africans, 4) mean number of household members, 5) proportion of households with school-aged children (aged ≤13 years), and 6) proportion of households that had experienced mortality of a child under five years old.

For each community-level variable, the median and interquartile range (IQR) are reported for the 93 study communities and by type of community: urban, peri-urban, and rural. Robust linear regression was used to test for associations between each community-level variable and community-level TB stigma (dependent variable). Interaction terms between each community-level variable and urban, peri-urban, or rural status were tested in the regression models using the Wald test. Stratified results are reported when p-value <0.20. All analyses were performed using Stata version 16.1 (College Station, Texas).

### Ethics statement

This study was approved by the University of Cape Town human research ethics committee (HREC Ref: 532/2020) and the Eastern Cape Provincial Research Office (Ref #: EC_202110_011). All study staff received training on Protection of Human Subjects and Good Clinical Practice. Data collection was done in a private location, when possible. Participants were compensated for their time with a food voucher.

### Patient and public involvement

We actively engaged with community members during the design of this portion of the study. We utilized workshops with individuals living in or familiar with the planned study areas to ensure community-informed demarcation of study communities. This was particularly valuable when no clear boundary existed (such as a major road). Our study team also includes a community liaison who met with Ward Counselors under whose jurisdiction the study communities fell. The Ward Counselors were informed of the purpose of the study and agreed to allow household recruitment in their areas. While they did not directly assist with participant recruitment, they did communicate to the public that a research study would be on-going in the area and assisted the field teams in addressing any challenges that arose. Once we have results identifying important social determinants impacting the TB care cascade, community-based engagement will occur to seek input and begin planning future interventions.

## RESULTS

Enrollment details for the 93 communities, grouped by their geographic areas, are reported in Supplemental Table S1. The median number of squares surveyed per community was four and ranged from three to eight, with rural areas requiring more squares to achieve the minimum target sample size per community. The median number of households surveyed was above the target of 24-36 per community, primarily driven by higher-than-expected recruitment. The median participation across a In total, 3,869 households were surveyed, with 18% coming from rural areas.

Household demographics varied by urban, peri-urban, or rural location (Table 1). Urban communities tended to have younger mean age, a lower proportion of individuals with matric (i.e., high school diploma), lower proportions of black South Africans, and lower prevalence of households ever experiencing TB or HIV. Meanwhile, rural communities were older, had more female HoHs, were all black South African, had more household members (including younger household members), experienced low under-five mortality, and had the highest prevalence of households ever experiencing TB or HIV. Peri-urban communities shared characteristics with both urban and rural communities. Like rural communities, peri-urban communities were mostly black South African, had a higher proportion of individuals with matric, and had experienced more household TB. Like urban communities, peri-urban communities had fewer household members, and more experiences of under-five mortality.

**Table 1.**
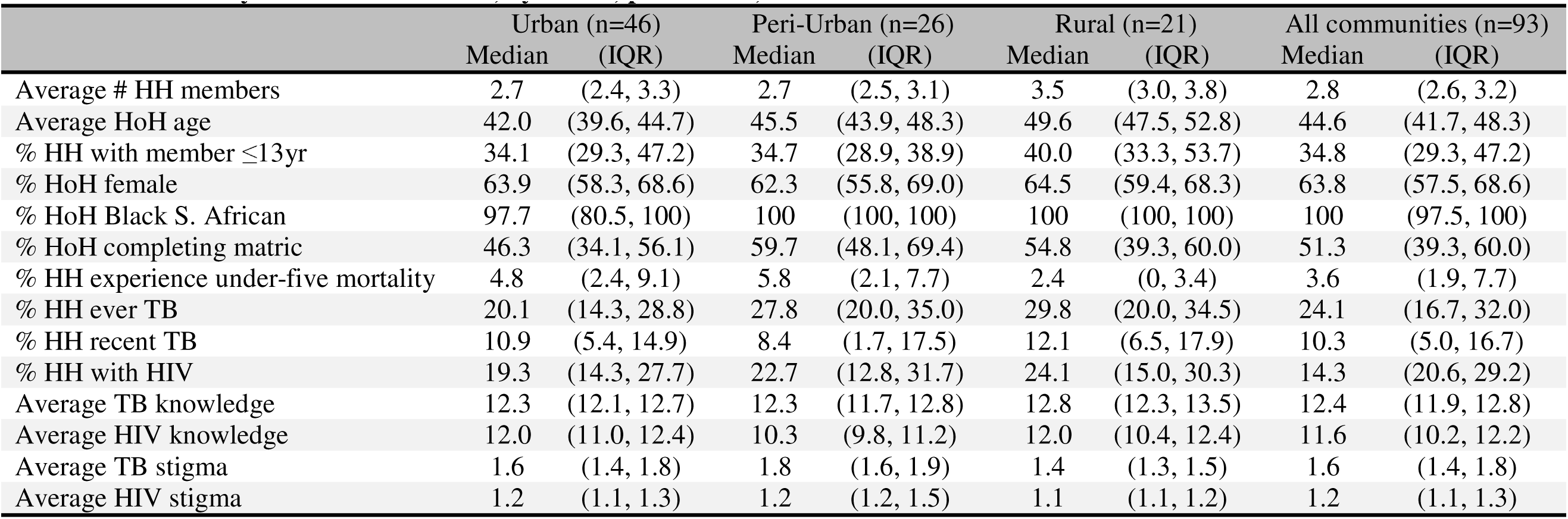
Community-level characteristics, by urban, peri-urban, and rural communities.

|  | Urban (n=46) |  | Peri-Urban (n=26) |  | Rural (n=21) |  | All communities (n=93) |  |
| --- | --- | --- | --- | --- | --- | --- | --- | --- |
|  | Median | (IQR) | Median | (IQR) | Median | (IQR) | Median | (IQR) |
| Average # HH members | 2.7 | (2.4, 3.3) | 2.7 | (2.5, 3.1) | 3.5 | (3.0, 3.8) | 2.8 | (2.6, 3.2) |
| Average HoH age | 42.0 | (39.6, 44.7) | 45.5 | (43.9, 48.3) | 49.6 | (47.5, 52.8) | 44.6 | (41.7, 48.3) |
| % HH with member ≤13yr | 34.1 | (29.3, 47.2) | 34.7 | (28.9, 38.9) | 40.0 | (33.3, 53.7) | 34.8 | (29.3, 47.2) |
| % HoH female | 63.9 | (58.3, 68.6) | 62.3 | (55.8, 69.0) | 64.5 | (59.4, 68.3) | 63.8 | (57.5, 68.6) |
| % HoH Black S. African | 97.7 | (80.5, 100) | 100 | (100, 100) | 100 | (100, 100) | 100 | (97.5, 100) |
| % HoH completing matric | 46.3 | (34.1, 56.1) | 59.7 | (48.1, 69.4) | 54.8 | (39.3, 60.0) | 51.3 | (39.3, 60.0) |
| % HH experience under-five mortality | 4.8 | (2.4, 9.1) | 5.8 | (2.1, 7.7) | 2.4 | (0, 3.4) | 3.6 | (1.9, 7.7) |
| % HH ever TB | 20.1 | (14.3, 28.8) | 27.8 | (20.0, 35.0) | 29.8 | (20.0, 34.5) | 24.1 | (16.7, 32.0) |
| % HH recent TB | 10.9 | (5.4, 14.9) | 8.4 | (1.7, 17.5) | 12.1 | (6.5, 17.9) | 10.3 | (5.0, 16.7) |
| % HH with HIV | 19.3 | (14.3, 27.7) | 22.7 | (12.8, 31.7) | 24.1 | (15.0, 30.3) | 14.3 | (20.6, 29.2) |
| Average TB knowledge | 12.3 | (12.1, 12.7) | 12.3 | (11.7, 12.8) | 12.8 | (12.3, 13.5) | 12.4 | (11.9, 12.8) |
| Average HIV knowledge | 12.0 | (11.0, 12.4) | 10.3 | (9.8, 11.2) | 12.0 | (10.4, 12.4) | 11.6 | (10.2, 12.2) |
| Average TB stigma | 1.6 | (1.4, 1.8) | 1.8 | (1.6, 1.9) | 1.4 | (1.3, 1.5) | 1.6 | (1.4, 1.8) |
| Average HIV stigma | 1.2 | (1.1, 1.3) | 1.2 | (1.2, 1.5) | 1.1 | (1.1, 1.2) | 1.2 | (1.1, 1.3) |

Across all study communities, the median community TB stigma score was 1.6, slightly higher than the midpoint of possible scores (Figure 2). Community stigma ranged from a low score of 1.0 (community members tended to disagree with TB stigma survey items) to a high score of 2.5 (community members tended to agree or strongly agree with TB stigma survey items), reflecting considerable variability in community attitudes. Compared to urban communities, rural communities had TB stigma scores 0.235 points lower (95% CI: -0.362, -0.108), while peri-urban communities had TB stigma scores that were 0.136 points higher (95% CI: 0.017, 0.254).

**Figure 2.**
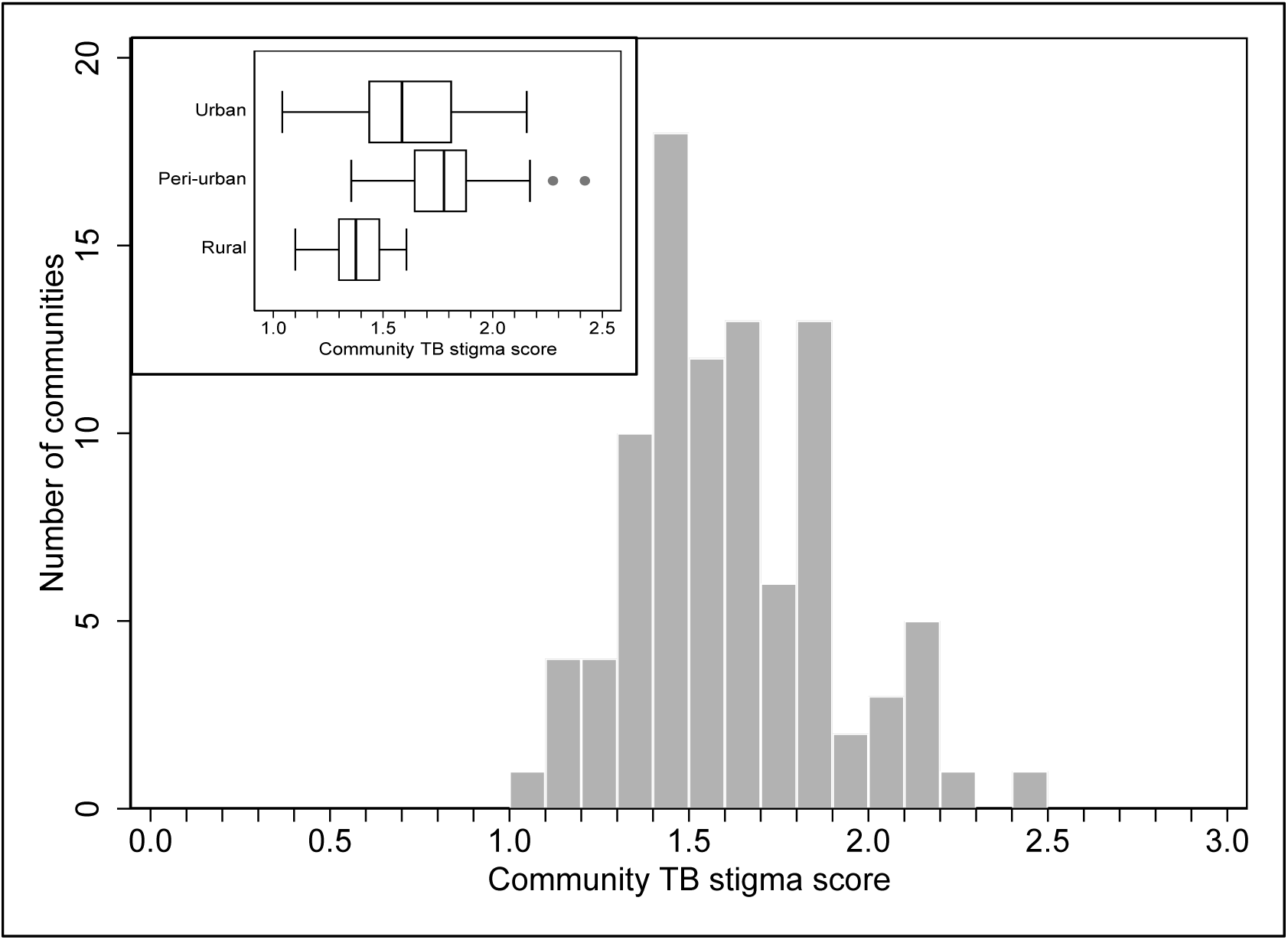
Distribution of community TB stigma scores across the 93 study communities (histogram) and by location (box plot inset)

Scatterplots of all community-level predictors and community TB stigma are presented in Supplemental Figures 2a through 2l) and robust regression results are presented in Table 2. There was a slight increase in TB stigma scores for communities with a higher proportion of households with TB in the past two years, but this was not statistically significant. Meanwhile, communities with better TB knowledge had lower TB stigma scores, though this was not significant after adjusting for study location. All p-values for interactions between community-level characteristics and urban, peri-urban, or rural status were non-significant (all P >0.25) except for HIV stigma (p=0.004). Stratified results by urban, peri-urban, and rural status can be found in Supplemental Table S2.

**Table 2.** Crude and adjusted robust regression results for community-level predictors of community TB stigma.

|  | Crude beta estimate | (95% Confidence Interval) | Adj.<sup>1</sup> beta estimate | (95% Confidence Interval) |
| --- | --- | --- | --- | --- |
| Average # HH members | **-0.141** | **(-0.234, -0.048)** | -0.041 | (-0.140, 0.058) |
| Average HoH age, per 10 years | **-0.147** | **(-0.268, -0.027)** | -0.063 | (-0.207, 0.082) |
| % HH with member ≤13yr, per 10% | **-0.049** | **(-0.097, -0.000)** | -0.016 | (-0.060, 0.029) |
| % HoH female, per 10% | -0.026 | (-0.102, 0.049) | -0.010 | (-0.076, 0.055) |
| % HoH completing matric, per 10% | -0.004 | (-0.043, 0.036) | -0.025 | (-0.061, 0.010) |
| % HH experience U5 mortality, per 10% | 0.067 | (-0.061, 0.194) | -0.032 | (-0.150, 0.085) |
| % HH ever having TB, per 10% | 0.000 | (-0.056, 0.056) | 0.001 | (-0.050, 0.052) |
| % HH recent TB, per 10% | 0.033 | (-0.047, 0.113) | 0.039 | (-0.029, 0.107) |
| % HH with HIV, per 10% | -0.001 | (-0.060, 0.057) | 0.003 | (-0.047, 0.053) |
| Community TB knowledge score | **-0.086** | **(-0.166, -0.007)** | -0.036 | (-0.110, 0.038) |
| Community HIV knowledge score | **-0.052** | **(-0.097, -0.007)** | -0.034 | (-0.076, 0.009) |

<sup>1</sup> Adjusted for urban, peri-urban, or rural status

As with community TB knowledge, some community characteristics were initially associated with lower community TB stigma, yet all were attenuated after adjusting for urban, peri-urban, or rural location. There were, however, two exceptions. There was a strong association for community HIV stigma and community TB stigma scores, with the magnitude of association differing by community type (Figure 3): Both urban and rural communities had a nearly one-point increase in TB stigma score for each one-point increase in HIV stigma score, while in peri-urban communities the association was substantially attenuated, though still significant. Urban communities with a higher proportion of black South Africans also showed lower community TB stigma scores (Supplemental Table S2). Study participants from peri-urban and rural communities were all black South African so no analyses were done for those communities.

**Figure 3.**
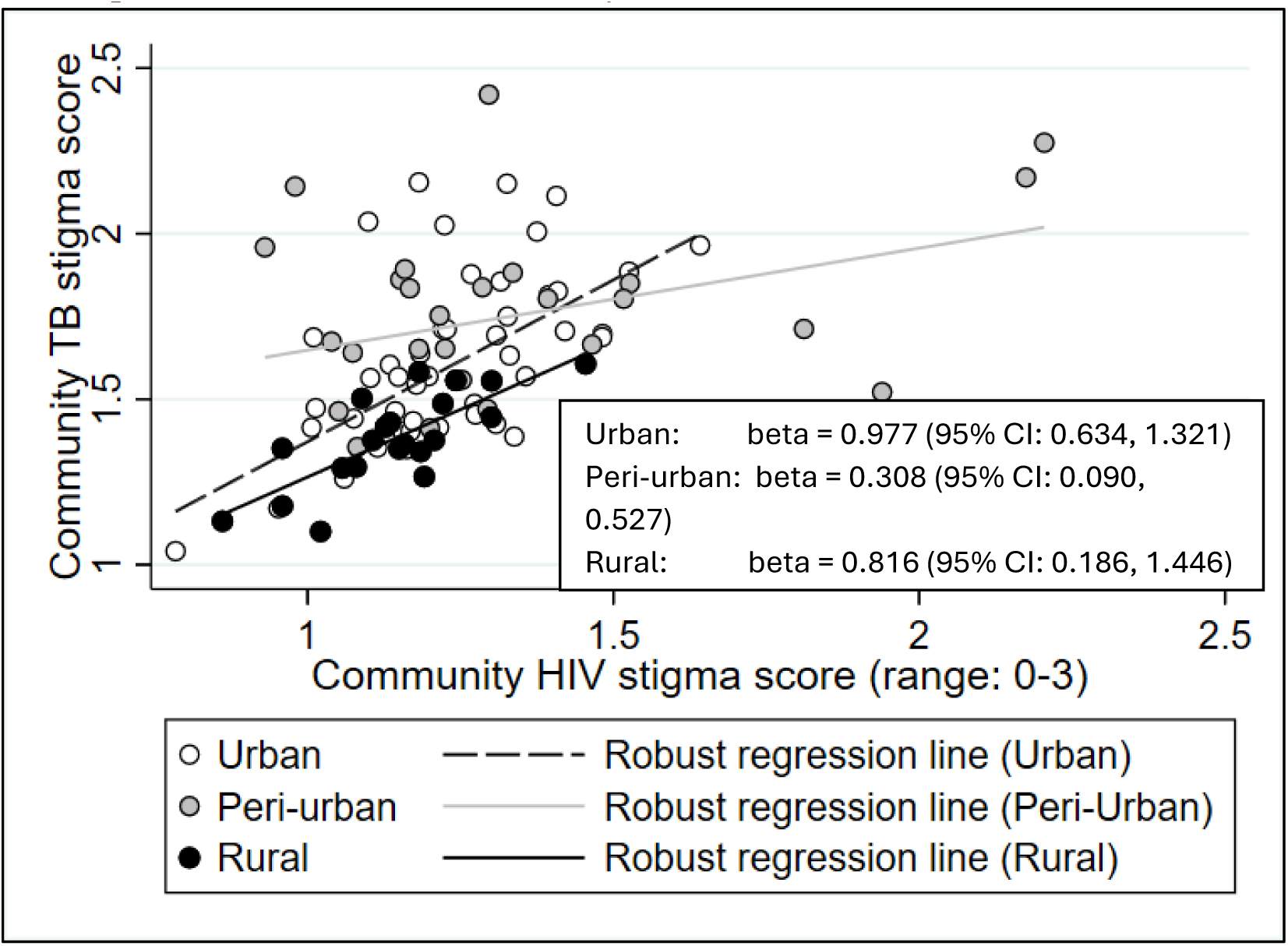
Associations between community TB stigma scores and community HIV stigma scores, by urban, peri-urban, and rural community status.

## DISCUSSION

The primary objective of this study was to obtain community-level estimates of TB stigma for use in subsequent multi-level analyses of stigma and engagement with the TB care continuum. We surveyed 93 communities across urban, peri-urban, and rural locations and found considerable variability in community TB stigma levels as well as prevalence of households experiencing TB and HIV. Rural communities generally had low levels of TB stigma while peri-urban communities had higher levels of TB stigma. Interesting, some urban communities reported low TB stigma while others reported higher stigma. A similar but less pronounced trend was seen for HIV stigma. Across all communities, those with higher HIV stigma also reported higher TB stigma. Community household TB and HIV prevalence were not found to be associated with TB stigma.

We are not aware of many studies that have evaluated community or regional variability in TB stigma with such a large sample of communities, let alone investigate factors that may account for such variability. Bond et al. reported on the proportion of households that believed TB is punishment for bad behavior across 24 communities in Zambia and South Africa^67^, with communities ranging from 1% to 32% of households endorsing blame. On average, however, Zambian communities had a higher prevalence of blame than South African communities. The authors suggest this may be due to South Africa having a higher prevalence of TB and therefore more familiarity with TB. However, they do not discuss what may account for community variability within each country. Similarly, Datiko et al. administered a TB scale to community members across regions in Ethiopia but did not investigate demographic or other factors that might explain the observed regional variation in TB stigma scores.^51^

Utilizing Ghana Demographic and Health Survey (DHS) data, another study analyzed responses to the question “If a member of your family got TB, would you want it to remain a secret or not?” as a proxy for TB stigma.^49^ The regional variability in TB secrecy found in the Ghanan study was attributed to cultural variations in how TB disease is perceived. Dormechele et al. evaluated the same TB secrecy question in DHS data from seven Sub-Saharan Africa countries.^53^ While there was variability in responses to this question across countries, they did not investigate potential explanatory factors. Finally, Rood et al. utilized the same question to conduct a multi-level analysis of DHS data from 13 countries (7 in Africa).^50^ They found that in countries with a higher national TB incidence rate there was less intent to keep a TB diagnosis secret. In contrast, countries with a higher levels of TB knowledge and higher HIV prevalence had more intent to keep a TB diagnosis a secret. In their conclusion, the authors focus on HIV as the driving factor behind TB stigma, noting that TB infection often remains a marker of HIV and therefore negative attitudes about HIV are imposed on people with TB or at risk for TB. Our study did not find any association between community TB stigma or TB knowledge and the prevalence of households in the community that had experienced TB (ever or in the most recent two years). This could be due to the use of different measures for stigma and knowledge or our ecologic study design while Rood et al. evaluated individual TB stigma through a multi-level design. Like Rood et al., however, our study does report a strong association between higher community HIV stigma levels and higher community TB stigma. This is not surprising given the well-documented relationship between TB and HIV. HIV is the most common risk factor for TB disease and, in South Africa, ∼54% of people diagnosed with TB have HIV co-infection.^2^ This influences the social perception of TB which often conflate having TB with also having HIV. As a result, people with TB bear the burden of historic societal attitudes about TB in addition to stigmatizing attitudes related to HIV.^13, 68^ Prior studies that have measured both TB and HIV stigma report a strong correlation between the two among community members and/or individuals with TB^52, 60^, while individuals co-infected with TB and HIV also report higher stigma than those who are not co-infected.^60, 69^ Of particular interest in our study, however, was that the strength of the relationship between TB and HIV stigma was greatly attenuated among peri-urban communities. We are not aware of any other studies that have considered an urban/rural spectrum instead of a strict dichotomy.

Lastly, we found a significant association between TB stigma and a community being urban, peri-urban, or rural; rural communities had the lowest TB stigma scores while peri-urban communities had the highest. While urban and rural differences in stigma are often discussed, few studies have evaluated this. Studies that have focused on an urban/rural status have done so as an individual-level characteristic, not a characteristic of communities, and they report conflicting results. In contrast to our findings, a study from Nigeria found that urban community members reported higher TB stigma than rural individuals^70^, while studies from Ethiopia and South Africa have reported no association with TB stigma.^51, 52^ Studies utilizing DHS data on intent to keep a TB diagnosis secret are also inconclusive.^49, 53, 71^

As with all population surveys, various limitations should be considered. First, it is possible that our sample is not representative of the target population. We utilized a single-stage, geographic cluster sampling process that balanced the need for representative sampling with the logistical challenges in the field. Our study communities included both urban and rural settings where reliable enumeration of households was not available. As a result, individual household selection utilizing probability proportional to size, as is done in many two-stage survey sampling designs, was not feasible. We did not rely on single clusters, but rather ensured three or more clusters were surveyed per community. This ensured a more representative population. Second, we surveyed the HoH for each household sampled. We believed that the HoH would have the most complete knowledge about that household, and that their attitudes and perceptions would influence the attitudes and perceptions of other household members. However, if TB stigma differed within households, our representation of community-level TB stigma may not be fully accurate. Third, all data were self-reported by participants. This means we cannot rule out social desirability or poor recall. Nevertheless, we did use validated measures of stigma that capture forms of stigma while minimizing social desirability bias. Knowledge of a family member having TB is likely accurate, though non-disclosure and under-reporting of household health characteristics may also have been possible. Fourth, selection bias due to non-participation is always a concern in household surveys. While participation was >70% in nearly all study communities, community stigma scores would be biased if those who refused to participate (perhaps due to stigma) or were not home during surveying periods differed in their attitudes.

This study also has major strengths, making it an important contribution to the scientific literature on TB stigma. First, to our knowledge, this is the only study documenting variability across many communities representing urban, peri-urban, and rural areas, thus making it more informative than studies focused only on regional or national data. Moreover, it was conducted in a setting with a very high TB burden that also suffers from many of the structural drivers of TB. Although focused on the Eastern Cape Province in South Africa, our findings may be relevant for other high TB/HIV burden areas, including both urban and rural. Second, many ecologic or multi-level studies traditionally rely on census units for their sampling and data collection. Our engagement with community members to more meaningfully demarcate smaller community areas within clinic catchment areas allowed for more meaningful division and variations between communities.

## CONCLUSION

Considering our study findings, urban versus rural status may be better viewed as a potential moderator of other drivers of TB stigma, rather than a driver in and of itself, as it is likely to be a proxy for other drivers of stigma. Aside from HIV stigma, we did not identify demographic or TB-related community-level factors associated with TB stigma. Stigma is a complex social process and there may be many other factors shaping TB stigma at the community level. Our on-going research aims to identify additional community-level drivers that can inform TB stigma interventions. Implementation of stigma interventions should consider the urban/rural spectrum when deciding how narrowly (urban areas only vs. rural areas only) or broadly (across the urban/rural spectrum) to implement the intervention.

## ACKNOWLEDGMENTS

We would like to thank all community members that participated in the survey; Freedom Mukomana, Mbasa Njomane, and Kuhle Fiphaza for development and maintenance of the REDCap project database; and the many field staff who carried out the household surveys across the 93 study communities.

## DATA AVAILABILITY

All data produced in the present study are available upon reasonable request to the corresponding author

## AUTHOR CONTRIBUTIONS

AMK helped with the conception of the overall project, acquisition of grant funds, design and selection of data collection procedures and measures, and carried out the primary analyses. He was primarily responsible for drafting the manuscript

DO helped develop study procedures and oversaw all aspects of study implementation including data collection, reporting, and training and quality control; he also helped select data collection measures and aided in drafting the manuscript

NS and LM helped define boundaries of study communities and oversaw field data collection teams

EF assisted with selecting areas for surveying and developing maps to aide the field data collection teams

KP assisted with sampling design and statistical analysis plan

AD helped with the conception of the overall project and drafting the manuscript

NN helped to develop study procedures for data collection, consultation and engagement with study clinics

AMM helped with the conception of the overall project, acquisition of grant funds, and aided in drafting the manuscript

All authors provided critical input on manuscript text and/or interpretation of results at various steps of the manuscript writing process. All authors have seen and approved of the final version.

