## Supplemental tables and figures for "Community variability in TB-related stigma in South Africa: an ecologic analysis from the MISSED TB Outcomes Study"

**Figure S1. Buffalo City Metropolitan Municipality (red dotted line) and location of urban (red boxes), peri-urban (green), and rural (purple) sites for the MISSED TB Outcomes study.**

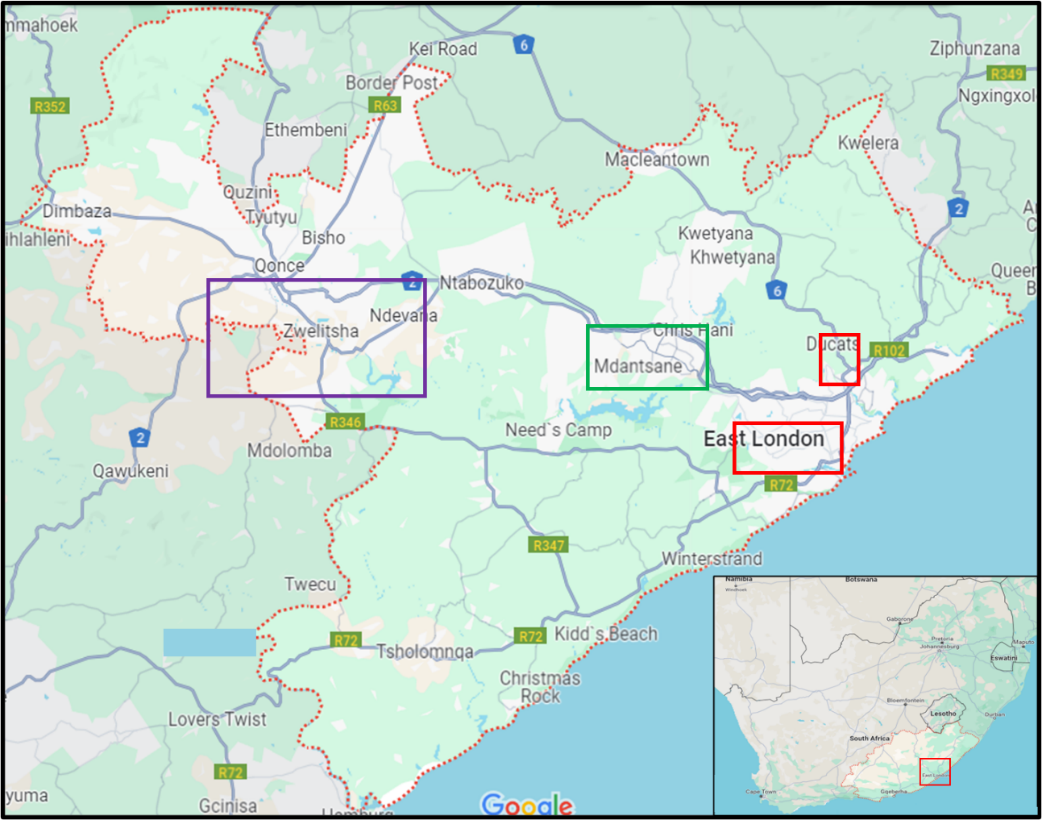

| **Table S1. Summary of community household enrollment** | | | | | |
| --- | --- | --- | --- | --- | --- |
|  | Number of communities | Median surveyed squares (IQR) | Median HH enrolled (IQR) | Median HH participation (IQR) | Total HH enrolled |
| Urban |  |  |  |  |  |
| Buffalo Flats | 7 | 4 (3, 6) | 42 (39, 63) | 75% (72%, 79%) | 328 |
| Ducats & Nompumelelo | 6 | 3 (3, 5) | 42 (33, 47) | 79% (75%, 88%) | 253 |
| Duncan Village | 25 | 4 (3, 4) | 38 (33, 41) | 93% (88%, 95%) | 941 |
| Scenery Park | 8 | 4 (3, 4) | 57 (43, 73) | 92% (86%, 96%) | 465 |
| Peri-Urban |  |  |  |  |  |
| Mdantsane | 26 | 4 (3, 5) | 44 (39, 50) | 87% (79%, 97%) | 1190 |
| Rural |  |  |  |  |  |
| Ndevana & Ginsberg | 10 | 5 (5, 7) | 32 (29, 40) | 78% (68%, 100%) | 346 |
| Zwelitsha | 11 | 6 (6, 7) | 30 (29, 33) | 63% (57%, 91%) | 346 |
| **TOTAL** | **93** | **4 (3, 6)** | **40 (33, 46)** | **87% (76%, 94%)** | **3869** |

Figure S2a. Scatterplot of community TB stigma scores and community average number of household members, by urban, peri-urban, and rural status.

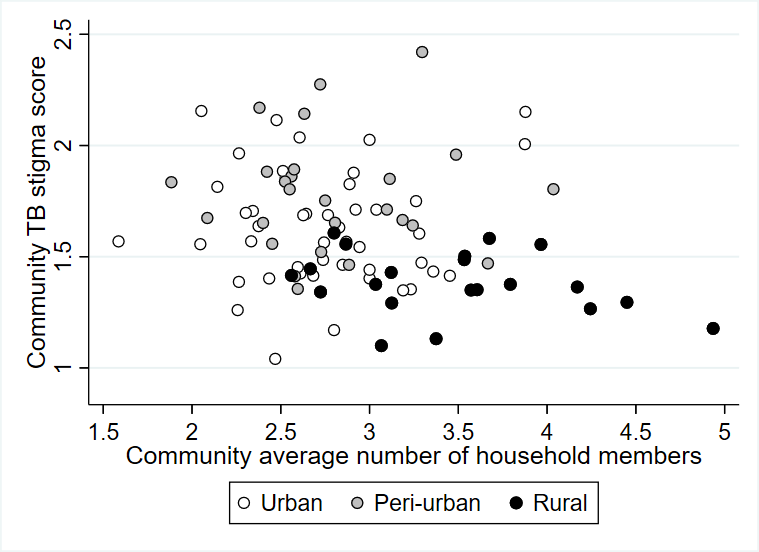

Figure S2c. Scatterplot of community TB stigma scores and community proportion of household members ≤13 years old, by urban, peri-urban, and rural status.

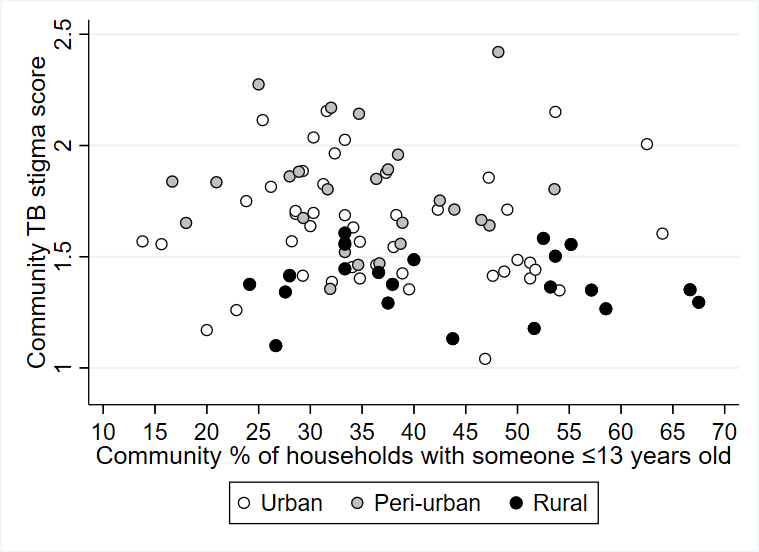

Figure S2b. Scatterplot of community TB stigma scores and community average number of household members, by urban, peri-urban, and rural status.

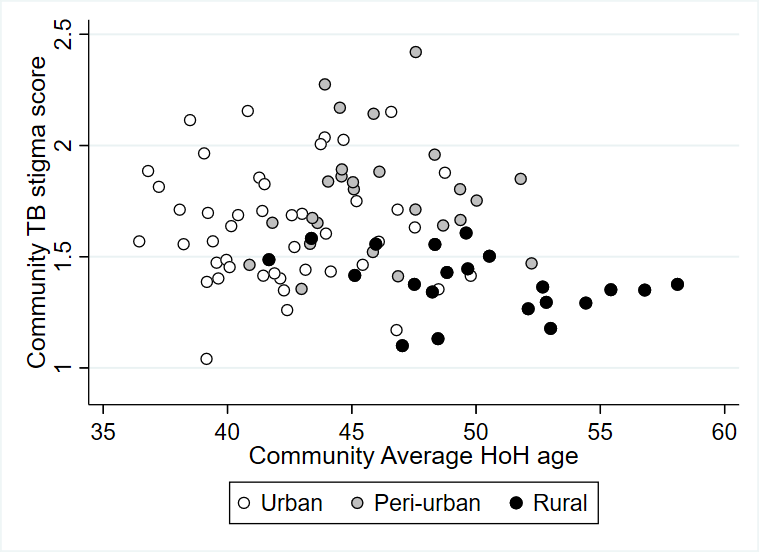

Figure S2d. Scatterplot of community TB stigma scores and community proportion of female heads of household, by urban, peri-urban, and rural status.

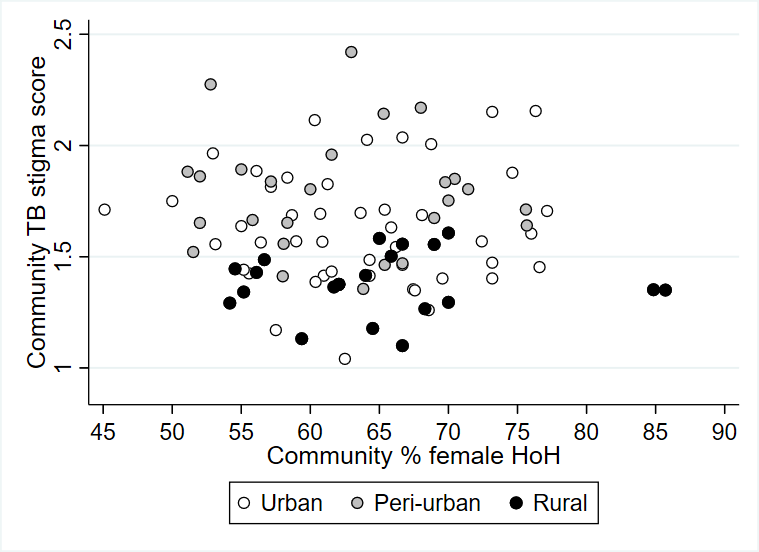

Figure S2e. Scatterplot of community TB stigma scores and community proportion of black South African heads of household in urban communities

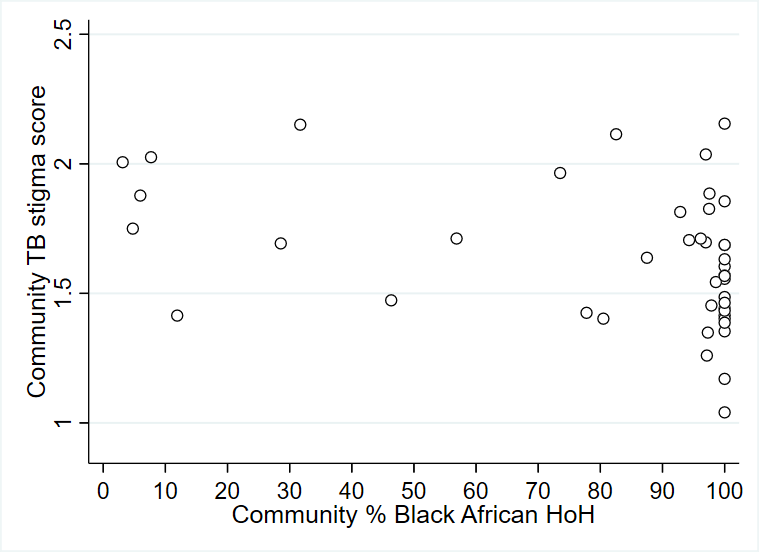

Figure S2g. Scatterplot of community TB stigma scores and community proportion of households having recent TB, by urban, peri-urban, and rural status.

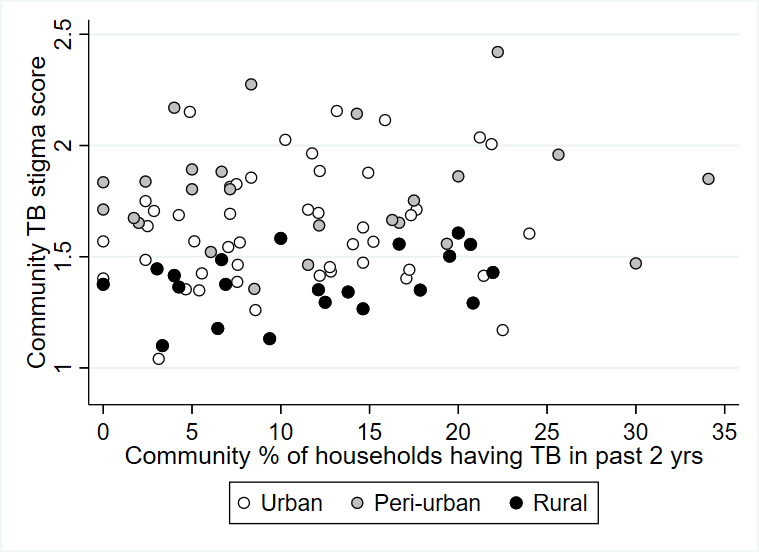

Figure S2f. Scatterplot of community TB stigma scores and community proportion of heads of household completing matric, by urban, peri-urban, and rural status.

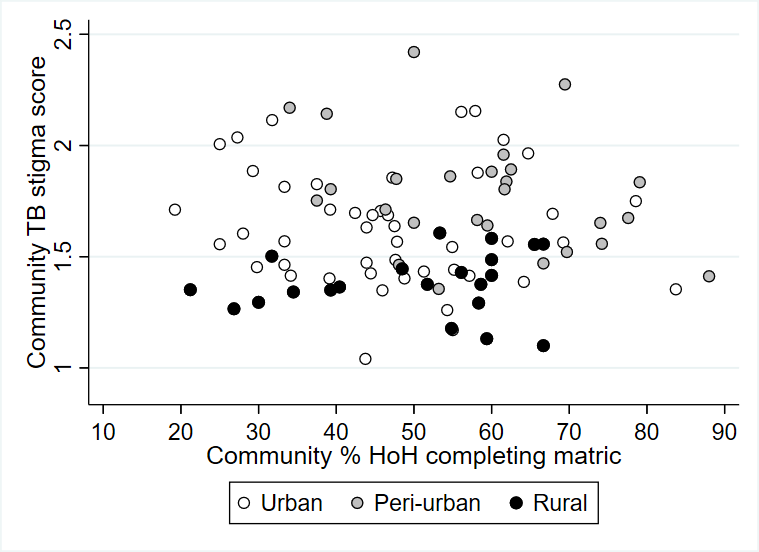

Figure S2h. Scatterplot of community TB stigma scores and community proportion of households ever having TB, by urban, peri-urban, and rural status.

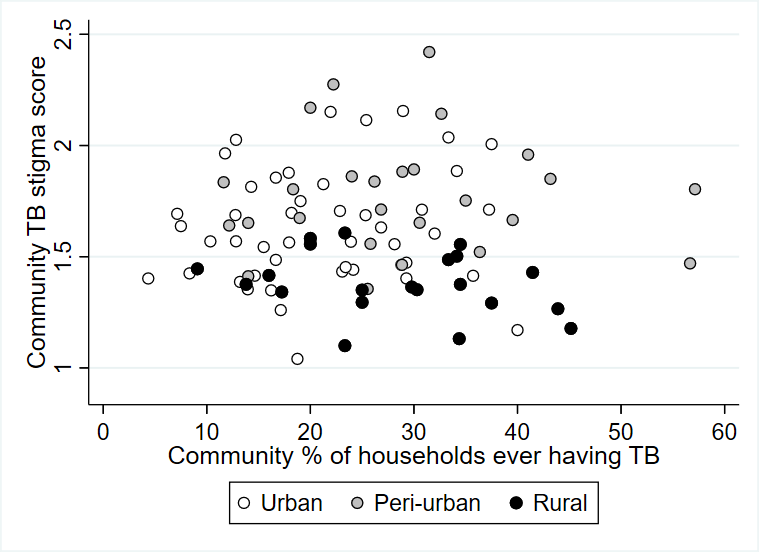

Figure S2i. Scatterplot of community TB stigma scores and community proportion of households living with HIV, by urban, peri-urban, and rural status.

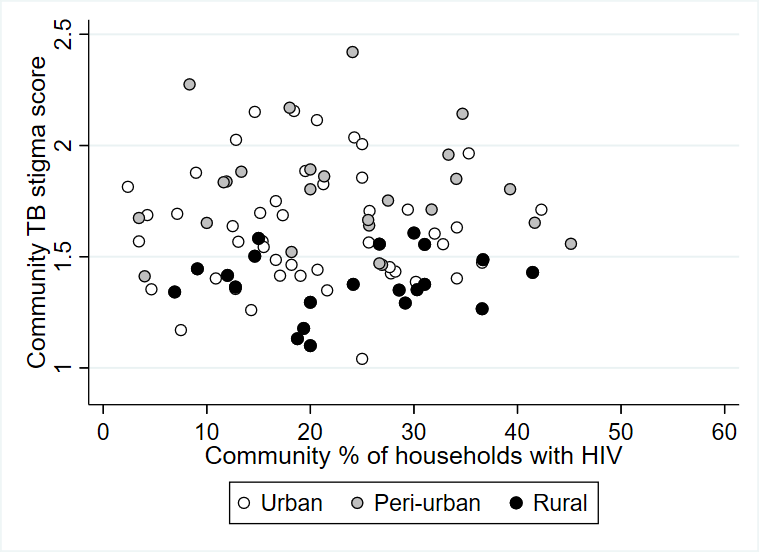

Figure S2k. Scatterplot of community TB stigma scores and community average TB knowledge score, by urban, peri-urban, and rural status.

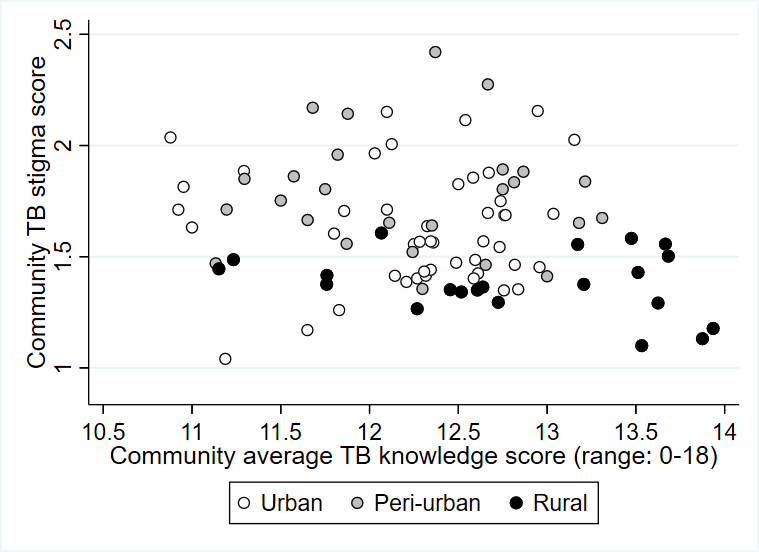

Figure S2j. Scatterplot of community TB stigma scores and community proportion of households with under-five mortality, by urban, peri-urban, and rural status.

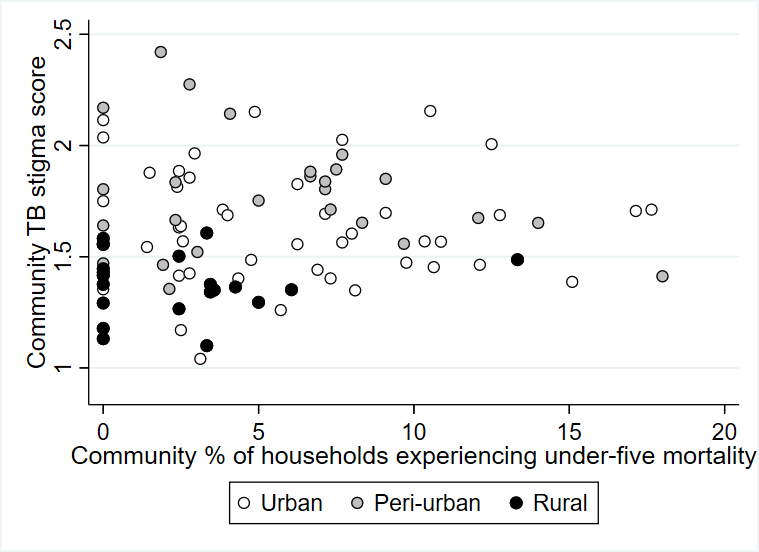

Figure S2l. Scatterplot of community TB stigma scores and community average HIV knowledge score, by urban, peri-urban, and rural status.

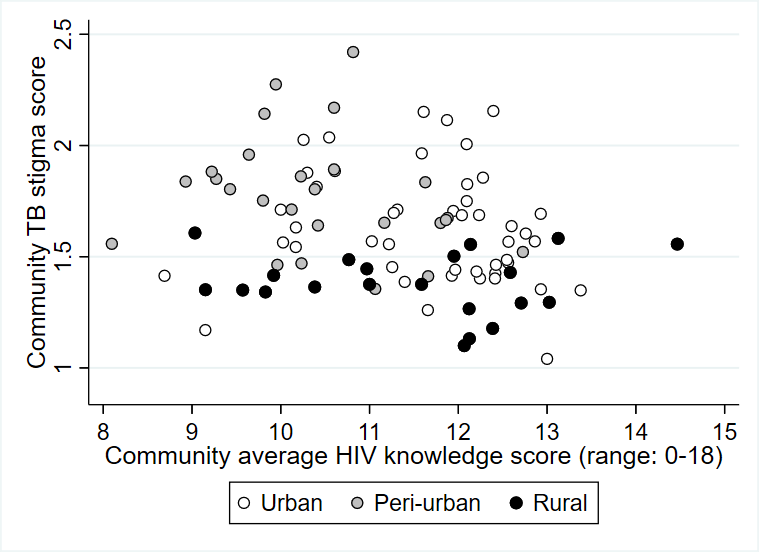

| **Table S2. Robust regression results for community-level predictors of community TB stigma, by urban, peri-urban, and rural status** | | | | | | | |
| --- | --- | --- | --- | --- | --- | --- | --- |
|  | Urban communities (n=46) | | Peri-urban communities (n=26) | | Rural communities (n=21) | |  |
|  | beta estimate | (95% CI) | beta estimate | (95% CI) | beta estimate | (95% CI) | Interaction  p-value |
| Average number of HH members | -0.008 | (-0.167, 0.150) | -0.029 | (-0.233, 0.174) | -0.067 | (-0.240, 0.107) | 0.88 |
| Average HoH age, per 10 years | -0.068 | (-0.283, 0.146) | 0.059 | (-0.270, 0.388) | -0.117 | (-0.371, 0.136) | 0.70 |
| % HH with member ≤13yr, per 10% | -0.022 | (-0.086, 0.041) | -0.017 | (-0.123, 0.090) | -0.007 | (-0.089, 0.075) | 0.96 |
| % HoH Black South African, per 10% | **-0.032** | **(-0.057, -0.008)** | n/a |  | n/a |  | n/a |
| % HoH female, per 10% | -0.023 | (-0.121, 0.074) | -0.007 | (-0.136, 0.122) | 0.002 | (-0.128, 0.133) | 0.95 |
| % HoH completing matric, per 10% | -0.024 | (-0.073, 0.024) | -0.067 | (-0.135, 0.0002) | 0.016 | (-0.059, 0.091) | 0.26 |
| % HH experiencing under-five mortality, per 10% | -0.006 | (-0.163, 0.150) | -0.116 | (-0.332, 0.100) | 0.014 | (-0.330, 0.358) | 0.68 |
| % HH ever having TB, per 10% | 0.037 | (-0.044, 0.118) | 0.0004 | (-0.081, 0.082) | -0.041 | (-0.150, 0.067) | 0.51 |
| % HH recent TB, per 10% | 0.060 | (-0.055, 0.174) | 0.009 | (-0.095, 0.113) | 0.068 | (-0.090, 0.226) | 0.74 |
| % HH with HIV, per 10% | -0.006 | (-0.082, 0.071) | 0.006 | (-0.081, 0.092) | 0.016 | (-0.098, 0.130) | 0.95 |
| Community TB knowledge score | -0.045 | (-0.168, 0.078) | -0.017 | (-0.165, 0.130) | -0.044 | (-0.169, 0.082) | 0.95 |
| Community HIV knowledge score | -0.045 | (-0.110, 0.021) | -0.065 | (-0.153, 0.023) | -0.005 | (-0.079, 0.068) | 0.55 |
| Community HIV stigma score | **0.977** | **(0.634, 1.321)** | 0.308 | (0.090, 0.527) | 0.816 | (0.186, 1.446) | 0.004 |

HH, Household
HoH, Head of household
n/a, not applicable: insufficient variation in % Black South African in the peri-urban and rural communities to perform analysis.
